# Coronavirus PPE: a positive pressure hood assembled from ubiquitous, low-cost materials

**DOI:** 10.1101/2020.04.14.20064808

**Authors:** Mark Crawford

**Affiliations:** University of Alberta

## Abstract

A positive pressure protective hood system was purposefully constructed only from materials commonly found worldwide, including bendable aluminum mesh, elastic head straps, velcro tape, a plastic sheet, a furnace filter and two computer central processing unit (CPU) cooling fans. The practical advantages of this system are that the materials are readily available in the inventories of most electronics and hardware outlets, ease of assembly (particularly if choosing to employ 3D printing for the fan enclosure and/or making several units at once with a defined workflow), and high probability of the materials being available in current or prospective personal protective equipment (PPE)-deplete regions. An experiment with identical fire detectors showed adequate inner isolation of the hood prototype from paper combustion particulates, which have a size range slightly smaller than putative coronavirus aerosols, for at least 90 seconds. The theoretical advantages of this system include significant reduction in healthcare provider exposure to coronavirus-containing respiratory fomites, respiratory droplets and aerosols (vs. traditional static masks and shields) during high risk procedures such as endotracheal intubation or routine care of an upright and coughing patient. Additionally, the assembly eliminates contact exposure to coronavirus fomites due to whole-head coverage from a hood system.

## Assembly

A fan platform was created from a semi-rigid, bendable material (bare aluminum mesh from orthopedic limb splints). Two extensions of the mesh were created on either side of the platform for stabilizing the device on the wearer’s head, as per a pair of glasses. Two Cooler Master Technology Inc.-brand 120 mm computer CPU fans (12v, static pressure 2.94 cmH_2_O, fan speed 2000 RPM) were placed on the platform side-by-side and assuring that airflow direction was downward, and then fully enclosed in wire mesh. The fans were then wired in parallel to a 12v battery source with an on/off switch, and the battery unit was mounted on the side of the aluminum platform with velcro tape (figure 1). Headlamp-style elastic head bands were used to provide a final adjustable fit, and foam padding was used within the unit to separate aluminum mesh material from the worker’s skin. A filter strip was cut from a home furnace filter rated 13 on the minimum efficiency reporting value (MERV) scale, and was double-taped into the plastic sheet (figures 2a,2b). This was the highest MERV-rated filter available at a local hardware store (a MERV rating of 13 is thought to filter particles in the range of 0.3-1 micrometers with 65% efficiency, 1-3 micrometers with 90% efficiency, and 3-10 micrometers with 98% efficiency)^1^. The filter was then double-taped to the top of the CPU fan enclosure. An unattached length of string was used to constrict the hood loosely at the level of the clavicle (figure 2c).

**Figure 1.**
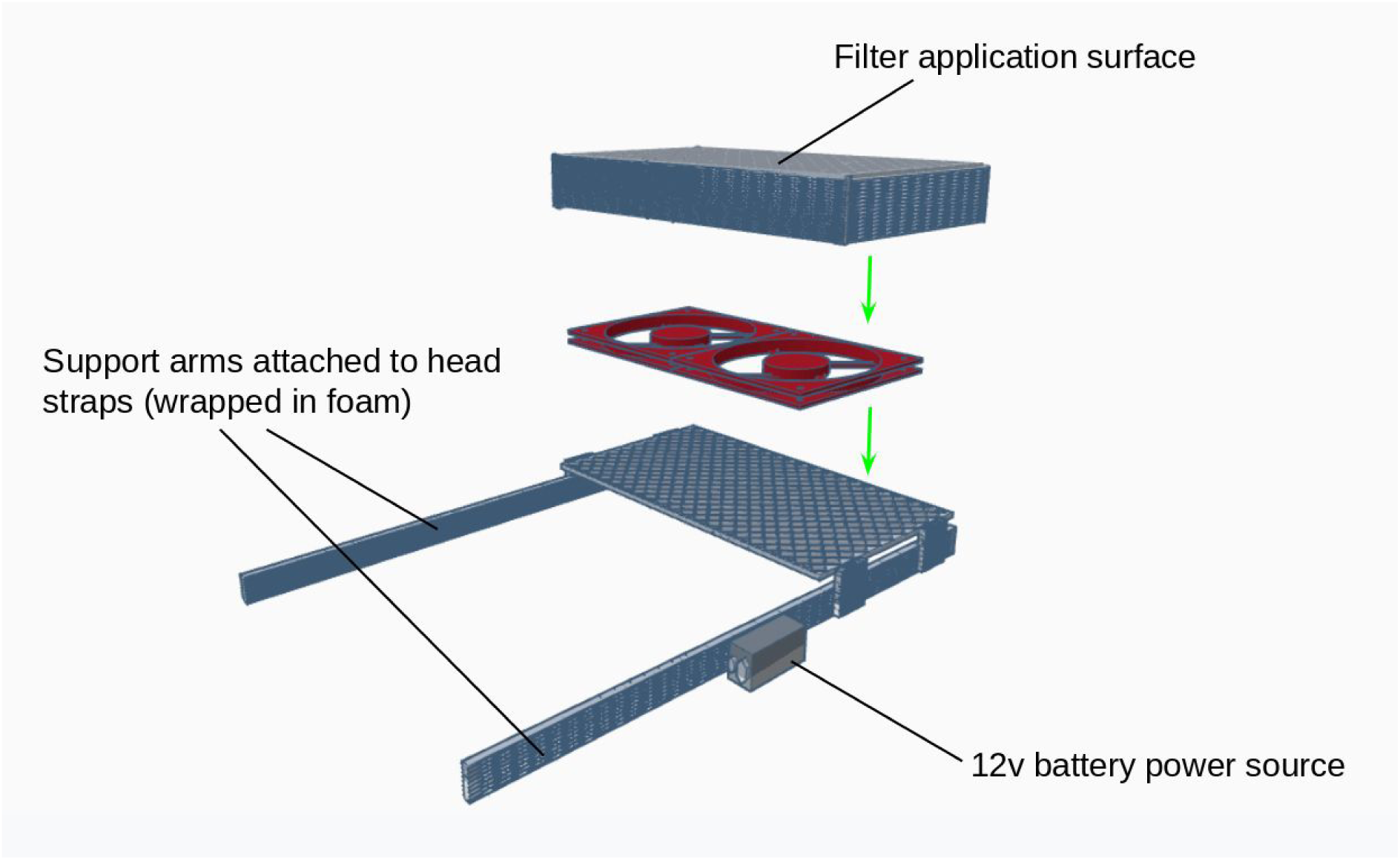
Assembly diagram of the fan enclosure. The two fans rest on the bottom of the enclosure, therefore airflow is directed downward with air being pulled through the filter material.

**Figure 2.**
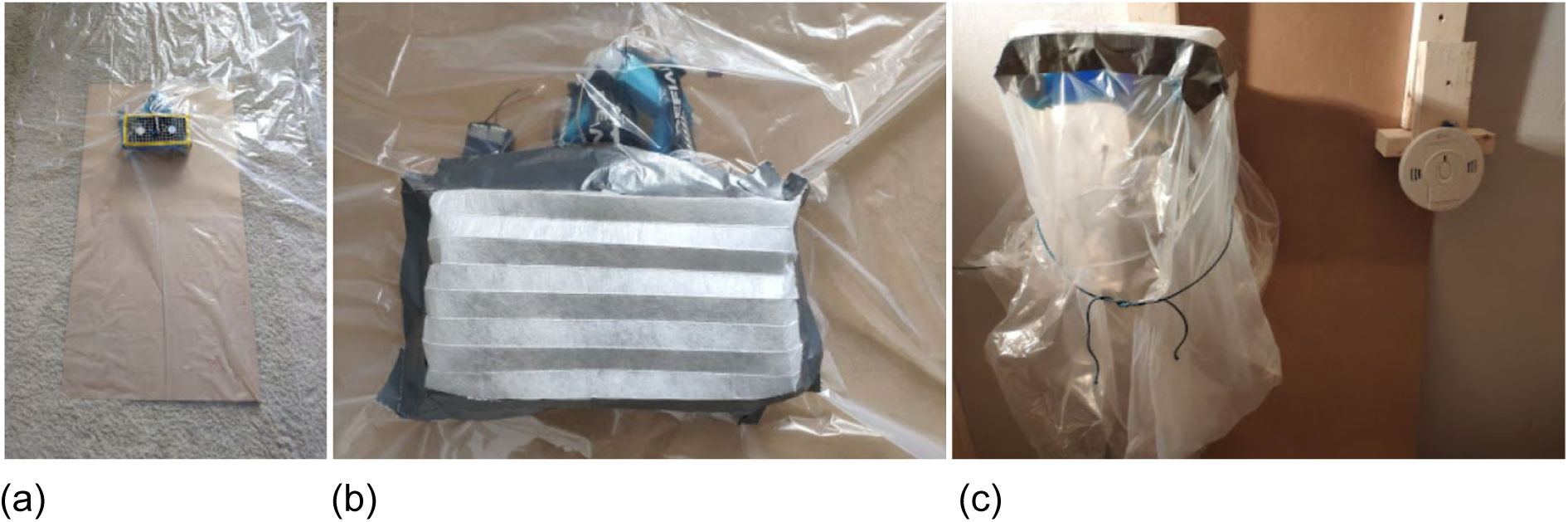
Double-taping the plastic sheet (a) and furnace filter (b) to the fan enclosure, and (c) the mounted hood assembly prior to smoke detector testing.

## Effectiveness

A small experiment was designed to determine the hood prototype’s ability to provide a protective environment from coronavirus-carrying particles. It was determined that paper combustion products (size range 0.04-0.08 micrometers) would be a reasonable surrogate for both respiratory droplets (0.6 −1000 micrometers) and the smallest coronavirus aerosols (0.06 - 1.40 micrometers)^2,3^. A small fire was started in a paper-filled glass bowl in a 4 m. × m. × 2.4 m. isolated room. Two identical, functioning fire detectors (Kidde Firex™ combination carbon monoxide & photoelectric smoke alarms (product number 900-0213CA, on battery power supply for experiment and therefore blinking every 30 s)) were mounted on a wall at the same height. The hood prototype was switched on and mounted over the left-sided fire detector, with the other detector remaining uncovered. After generating a large enough smoke plume for detection by the uncovered alarm, the fire detector inside the hood did not alarm during a smoke exposure period of 30 seconds. The experiment was repeated with the same filter in place and for 90 seconds of alarm time of the uncovered detector (with more paper burning in a larger steel container). In both scenarios, the fire detector inside the hood did not alarm.

## Practical points

- The prototype was worn by an adult male for 40 minutes continuously. There was continuous airflow to the head and face, and no noticed breathing difficulty while ambulating and on mild-moderate exertion. The movements required by a healthcare worker to conduct airway management or other routine procedures would not be hindered by the apparatus.
- To test the durability of these specific CPU fans operating at full speed (ie., not investigating battery life), the hood system was connected to a 12v adaptor power supply and remained running at full speed for a full 24 hours before being disconnected for storage.
- This hood system should be suitable for providing care to patients in need of high flow oxygen, as all electrical components are insulated and within a disposable plastic hood covering away from the bottom of the semi-constricted plastic sheet opening.
- A more rigid and visibility-enhancing clear vinyl sheet could be embedded into the plastic sheet if desired by the user.
- The contained and more expensive reusable portion could likely be cleaned with disinfectant wipes or a disinfectant spray.
- If the prospect of reusing the fan assembly is deemed unsafe by a health authority or individual user, the entire unit could be discarded at an estimated cost of CAD $50.17 / USD $35.62 / EURO €32.61. Single use of a commercial powered air-purifying respirator (PAPR) for one patient exposure can cost many multiples of this amount.
- As two axial fans in series can double the static pressure of the original fan area, it would be possible to add another layer/additional fan layers to the enclosure to optimize pull of air through the filter material (if weaker fans are only available in certain locales and/or if more filter material is deemed necessary).

## Context

The worldwide personal protective equipment (PPE) shortage during the Covid-19 pandemic has continued to endanger the lives of healthcare workers. Along with re-tooling factories, labour crowd sourcing and donations of stockpiled resources, novel assemblies of existing materials have the potential to reduce healthcare worker morbidity and mortality during the pandemic. This positive pressure hood system prototype may also serve to protect healthcare workers in established pre-pandemic low resource settings who will be experiencing greater PPE availability strain than healthcare systems in wealthier, higher-GDP jurisdictions. The enclosure of small, readily available fans to create a protective positive pressure hood system is the most important element of this submission, and modifications/improvements to the outlined prototype would certainly be possible.

Materials and CPU fans chosen may differ by region and the prototype has not been exposed to formal industrial/regulatory body testing. Exposure to fine particulate during initial prototype testing was limited to the materials available to an individual practicing social distancing, as well as the obvious risks of allowing a fire to smoulder in an enclosed space for an extended time period. If we are currently constructing PPE from non-traditional materials to protect healthcare workers during a pandemic, it will be the added responsibility of the builder to ensure that seams are well sealed and that the filter is properly positioned, with the final inspection by the user. An additional surgical or N95 mask could be worn within the hood at the wearer’s discretion or if available.

This hood system functions as a simple, low-technology PPE apparatus to create flow of filtered air during aerosolizing procedures and as a more complete face shield for fomite and droplet protection than N95 masks or open-bottom face shields. This apparatus would also reduce the risk of the healthcare provider inadvertently touching their exposed face, or of conjunctival/nasal/oral mucosal contact by fomites or aerosols. This assembly and disposable covering could be immediately translated to at-home and large scale production alike, with a portion of the global stock of small electronics cooling fans being redirected to protect healthcare providers during the coronavirus pandemic.

## Data Availability

There are no data outside of this manuscript

## Supplemental material

**Table S1.** Cost of materials sourced from local retailers in a Canadian city. Lower prices are currently available through online vendors if ordered in bulk.

| Item | CAD* | USD* | EURO* |
| --- | --- | --- | --- |
| Bare aluminum splints<br>(2)** | \$7.10 | \$5.04 | €4.62 |
| Elastic head straps (2)** | \$8.00 | \$5.68 | €5.20 |
| Master Cooler<br>Inc.-brand CPU fans 120<br>mm (2)** | \$18.00 | \$12.78 | €11.70 |
| AA batteries (8)** | \$6.67 | \$4.74 | €4.34 |
| Velcro tape (¾ inch x<br>18" strip)** | \$5.20 | \$3.69 | €3.38 |
| Approx. 300 mm x 150<br>mm piece of 3M Filtrete<br>MPR 2200 furnace filter<br>(gently flattened onto fan<br>cage)*** | \$2.00 | \$1.42 | €1.30 |
| Standard (see-through)<br>plastic construction<br>sheet*** | \$3.00 | \$2.13 | €1.95 |
| Electrical tape | \$0.20 | \$0.14 | €0.13 |
| **Total** | **\$50.17** | **\$35.62** | **€32.61** |
| **Total: re-usable**<br>**portion** | **\$45.17** | **\$32.07** | **€29.23** |
| **Total: disposable**<br>**portion** | **\$5.00** | **\$3.55** | **€3.25** |

\*Canadian dollars, US dollars and Euro currencies converted at time of submission.
\*\*Disposable portion of apparatus.
\*\*\*Theoretically reusable portion of apparatus.

**Supplemental video 1. Hood with fans on, camera facing cephalad from inside of hood**

**Supplemental video 2. Initial fire detector test: 30 seconds**

**Supplemental video 3. Second fire detector test, same filter as first: 90 seconds**

## References

1. Brown, K.W. et al. Reducing patientś exposures to asthma and allergy triggers in their homes: an evaluation of effectiveness of grades of forced air ventilation filters. J Asthma 51, 585–94 (2014).

2. He, X. et al. Laboratory evaluation of the particle size effect on the performance of an elastomeric half-mask respirator against ultrafine combustion particles. Ann Occup Hyg 57, 884–97 (2013).

3. Zhu, N. et al. A Novel Coronavirus from Patients with Pneumonia in China, 2019. N Engl J Med 382, 727–33 (2020).

